# Persistence of the T12 *Vibrio cholerae* O1 lineage in West Africa: Insights from a Regional Sequencing Workshop

**DOI:** 10.1101/2020.09.24.20199026

**Authors:** Eme Ekeng, Serges Tchatchouang, Blaise Akenji, Bassira Boubacar Issaka, Ifeoluwa Akintayo, Christopher Chukwu, Ibrahim Dan Dano, Sylvie Melingui, Ousmane Sani, Michael Oladotun Popoola, Ariane Nzouankeu, Yap Boum, Francisco J Luquero, Anthony Ahumibe, Dhamari Naidoo, Andrew S Azman, Justin Lessler, Shirlee Wohl

**Author notes:** authors contributed equally (Eme Ekeng, Serges Tchatchouang).

## Abstract

We sequenced 46 *Vibrio cholerae* isolates from Cameroon, Niger, and Nigeria, 37 of which were from 2018–2019. These sequences belong to the T12 lineage observed in the region since 2009, suggesting continuous transmission. Data were generated during a workshop in Nigeria, providing a model for future regionally coordinated surveillance efforts.

## Introduction

Molecular characterization of pandemic *Vibrio cholerae* has led to new insights about global cholera transmission and has highlighted the important role of transmission within and between Asia and Africa (1). Combining genomic and epidemiological data may allow us to better understand the dynamics of ongoing outbreaks, and advances in sequencing technology have recently made whole genome sequencing more feasible even in low-resource settings.

Although recent studies have used whole genome sequences of *V. cholerae* O1 to better understand the global movement patterns of seventh pandemic cholera in sub-Saharan Africa and elsewhere (1–4), regional and local dynamics in West Africa—which reports cholera regularly—are still poorly understood. Epidemiological data suggest outbreaks across this region may be connected (5–7), but the nature of recurring outbreaks in the region has yet to be understood. It is unclear if cases go unreported in years between outbreaks or if each represents a distinct pandemic *V. cholerae* introduction from outside the region. Discerning between these two transmission scenarios is important for the development of locally-adapted and effective cholera control and prevention strategies.

Here we focus on the 2018–2019 cholera outbreaks in three countries with endemic cholera—Cameroon, Niger, and Nigeria—to understand what *V. cholerae* sequence data from these outbreaks can tell us about transmission patterns within the region. These three countries all reported large numbers of cholera cases during this time period (2,066, 3,083, and 32,752, respectively) (8,9) after several years of reported low cholera incidence (**Fig 1A**). Broad sequencing of historical *V. cholerae* isolates included 45 sequences from Cameroon from 1970–2011, 15 sequences from Niger from 1970–2010, and 22 sequences from Nigeria from 1970–2014 (1,2). The specific lineage of *V. cholerae* circulating in each country, determined from the genomic data, provides important information about the movement of the disease.

**Figure 1.**
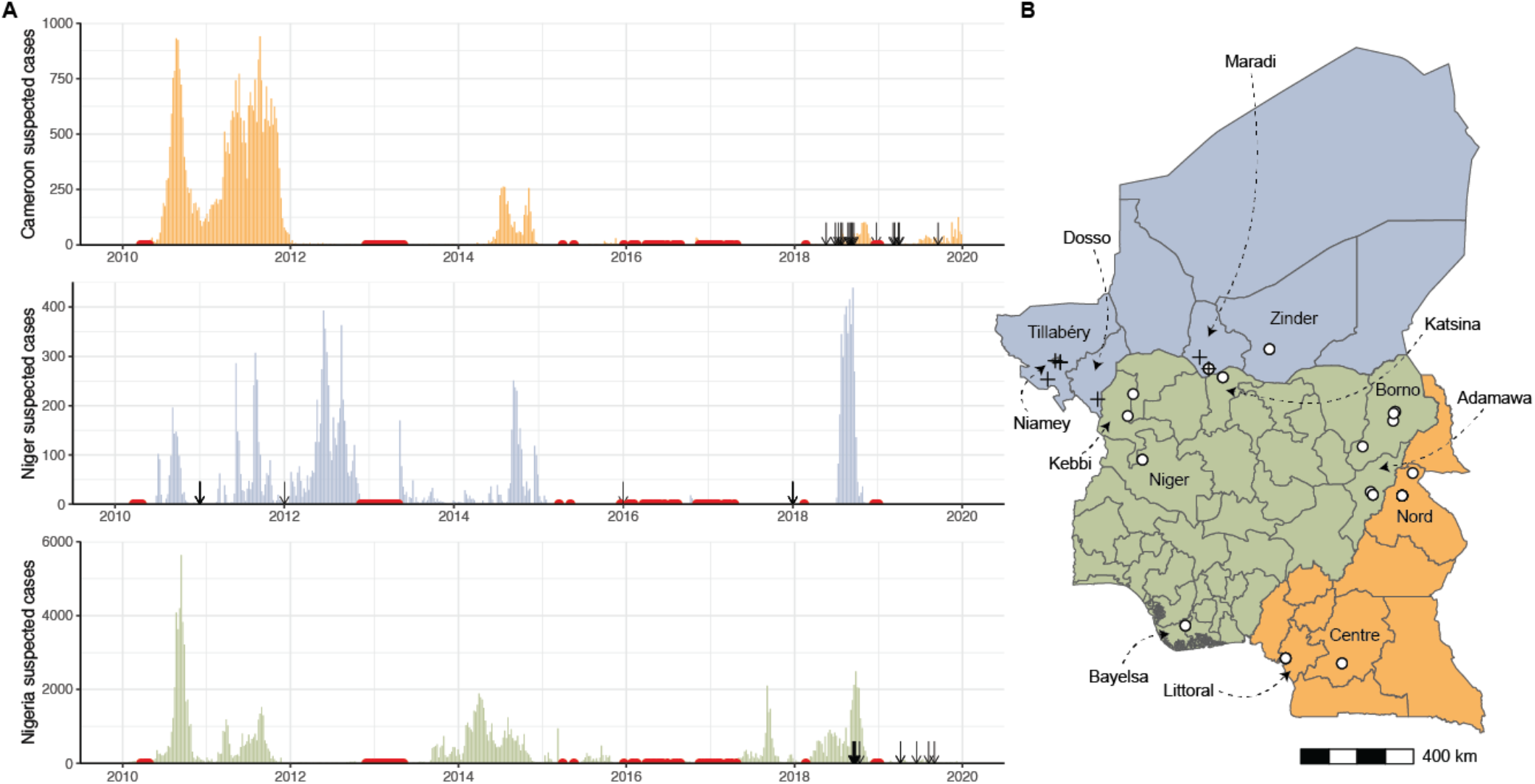
Cholera cases and sequenced isolates. (**A**) Weekly reported suspected cholera cases for Cameroon (orange), Niger (blue) and Nigeria (green) from 2010 through 2019 (8). Red points: weeks with no more than five suspected cases reported across all three countries. Arrows: collection dates of isolates sequenced. Collection dates provided as year only are plotted on January 1 of their given year. (**B**) Map of Cameroon, Niger, and Nigeria. Colors are as in (A). White points: location of sequenced isolates collected in 2018-2019. Black crosses: location of sequenced isolates collected prior to 2018. Two isolates with unknown sub-country locations are not shown.

Previous studies have shown that the T1 lineage was the predominant lineage circulating in Cameroon, Nigeria, and Nigeria in 1970, followed by the T7 and T9 lineages in the 1990s and early 2000s (1,2). Since 2009, only T12 has been observed in these countries, and this appears to be the predominant lineage in Western Africa and nearby countries in Central Africa. In any given year, the circulating *V. cholerae* lineage was the same in Cameroon, Niger, and Nigeria (1,2), providing additional evidence that outbreaks in these countries are connected. As five years had elapsed since the last published *V. cholerae* O1 genome in the region, we aimed to generate a contemporary snapshot of recent *V. cholerae* O1 outbreaks to determine if recent cases were due to a new introduction of the pathogen, and to gain new insights into regional cholera transmission dynamics and spread of antibiotic resistance.

## The Study

In October 2019, researchers from Cameroon, Niger, Nigeria, and the United States came together to discuss the specific cholera surveillance questions in these African countries and how whole genome sequencing can be used to address them. Simultaneously, researchers generated whole genome sequences from *V. cholerae* isolates collected in these countries and discussed plans for building a regional cholera genomics network to help inform cholera preparedness and outbreak response.

A key component of this workshop was bringing together detailed epidemiological data from the three countries and using this to select *V. cholerae* isolates for sequencing that captured the 2018–2019 outbreak over time and space. We also prioritized isolates from locations near the borders between the three countries, in order to increase the likelihood of capturing evidence of cross-border transmission. We selected 46 isolates from clinical and lab confirmed cases for sequencing (16 from Cameroon, 15 from Niger, 15 from Nigeria), 37 of which were from 2018–2019 cases (**Fig 1B, Appendix Table 1**).

We sequenced extracted DNA from these isolates on the Oxford Nanopore MinION and assembled genomes by aligning the resulting sequencing reads to the seventh pandemic O1 reference genome, strain N16961 (GenBank accession: AE003852/AE003853) (1,10). We also performed antimicrobial resistance (AMR) gene detection on these sequences and compared the results to previously published sequences.

Forty-four genomes produced reference-based assemblies (covering at least 95% of the N16961 reference at >100x coverage, **Appendix Table 2**). We aligned these genomes to 1280 previously-published *V. cholerae* O1 whole genome sequences (1,2,11,12) and generated a maximum likelihood tree from these data, which showed that all 44 isolates belong to the T12 lineage. Two sequenced isolates did not align to the reference (NGA_148_2019, NGA_252_2019), and we found that the reads did not map to the *wbe* or *wbf* gene clusters associated with the O1 and O139 serogroups, respectively (or the *ctxA* cholera toxin) (13). These results are concordant with laboratory testing performed in Nigeria, which designated these isolates as non-O1 *V. cholerae*. AMR profiles were similar to previously published sequences from this region, though we detected new mutations in genes (*gyrA* and *gyrB*) associated with quinolone resistance (see **Appendix** for detailed experimental methods and AMR results).

## Conclusions

We did not find evidence for a new introduction of *V. cholerae* O1 between 2014—the date of the last published isolate from these countries—and 2018. On the contrary, we found evidence for evolution of the bacterium within the region: all isolates from 2018–2019 (as well as the 2011–2016 isolates from Niger) fall within the T12 lineage, and the 2018–2019 sequences are part of a sub-clade containing 2014 and 2016 isolates from countries in West Africa (**Fig 2**). Within this sub-clade, the 2018–2019 sequences are distinct. In concordance with the epidemiologic data, the high similarity between sequences (including shared mutations in known AMR genes, see **Appendix Table 3** and **4**) from the three countries suggests that their outbreaks are linked.

**Figure 2.**
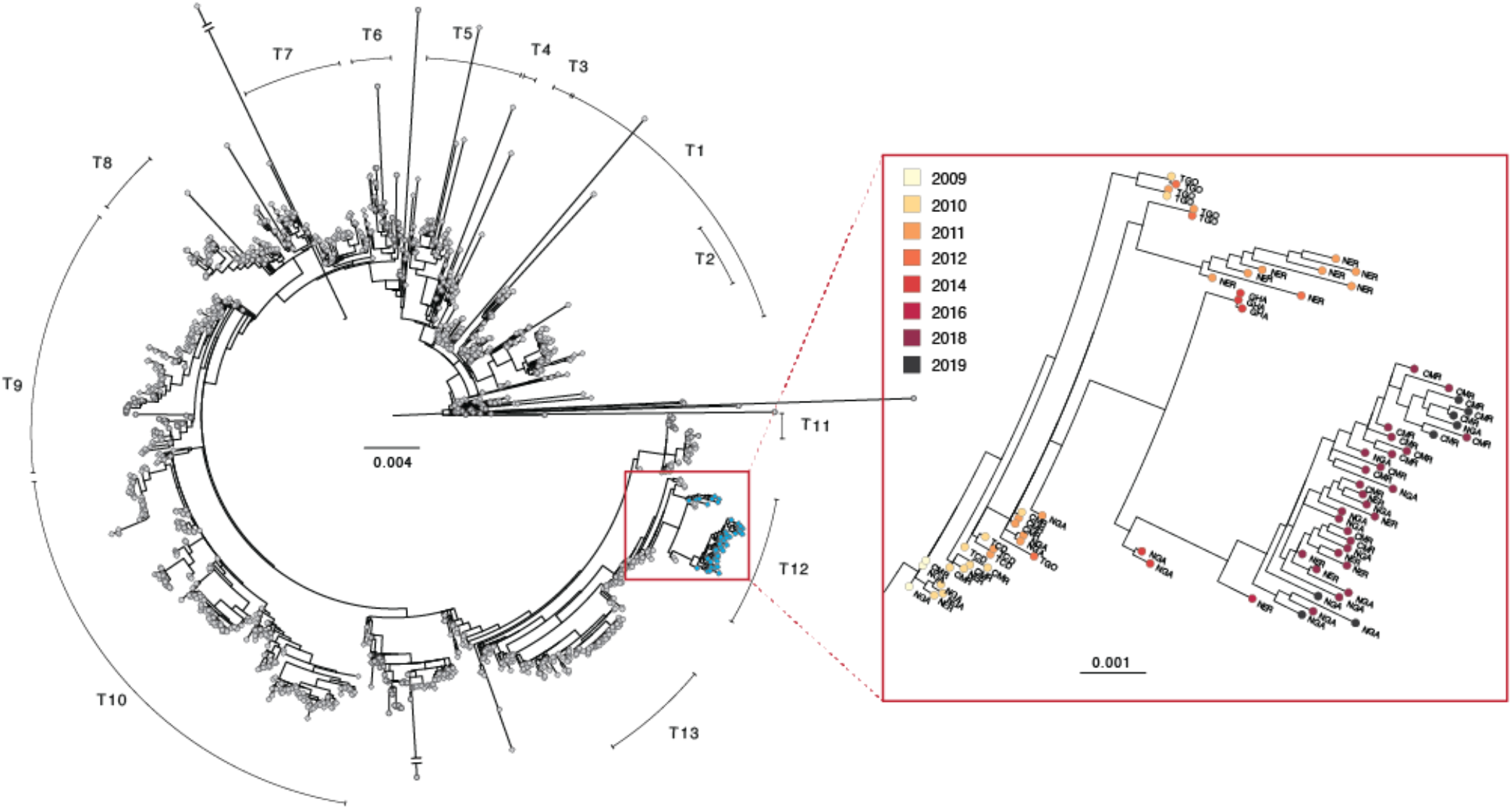
Phylogenetic tree of *V. cholerae* O1 sequences. Left: maximum likelihood tree of global *V. cholerae* isolates. Samples generated in this study are shown in blue. Right: zoom view of a portion of the T12 lineage containing *V. cholerae* genomes generated in this study. Country codes: TGO, Togo; NER, Niger; GHA, Ghana; CMR, Cameroon; NGA, Nigeria; TCD, Chad. Scale bar unit: nucleotide substitutions per site.

The phylogeny also suggests that nearby countries such as Ghana and Togo may have outbreaks connected to those in Cameroon, Niger, and Nigeria, though it is unclear just how far this cholera transmission region extends. As of this writing, there are no published *V. cholerae* O1 genomes from the nearby countries of Benin, Burkina Faso, or Mali, for example, from after 2010. Sequences from Central African Republic, South Sudan and Democratic Republic of the Congo from 2011–2018 suggest that the T10 lineage is predominant in Central Africa (2,12), but more recent data are needed to delineate the boundary between T10 and T12 circulation.

Even so, there is strong evidence that, at least in Cameroon, Niger, and Nigeria, T12 *V. cholerae* was present throughout 2009–2019, even though there were few reported cases in 2014–2017. This could be explained by underreporting or under-diagnosing of cases, movement of the same cholera lineage in and out of these countries during that time period, or an environmental reservoir of *V. cholerae* in the region (such as Lake Chad, though prior studies have provided evidence against this (14)) that is responsible for repeated reintroduction of the same lineage back into the population.

Expanded surveillance and additional sequencing data will be necessary to understand the interconnectedness of populations across countries and achieve global elimination goals such as those outlined in the GTFCC Roadmap. For example, limiting cross-border movement is an ineffective strategy in the presence of a natural reservoir responsible for repeated reintroduction. Additionally, sequencing data can be used to detect and track resistance to antibiotic treatments, and to identify potential new resistance-conferring mutations present in local isolates (15). A regional cholera surveillance network could facilitate efforts to gather isolates, build the local capacity needed to generate sequencing data, and ensure that these data are interpreted within the regional context. As a result of the workshop, scientists from Cameroon, Niger, and Nigeria have all been trained in *V. cholerae* sequencing with the Oxford Nanopore MinION platform; expanding sequencing knowledge and capacity beyond these three countries will make it easy to generate the data needed to control cholera outbreaks in the region.

## Data Availability

Raw data for all sequenced isolates are available at NCBI under BioProject accession: PRJNA616029.

## Acknowledgements

This project was made possible by the Nigeria Centre for Disease Control, who graciously hosted researchers at the National Reference Laboratory in Abuja, Nigeria for training and *V. cholerae* sequencing. We thank Aicha Omar for laboratory assistance and strain isolation, Alama Keita and the West Africa Cholera Platform for sharing incidence data, David Mohr for laboratory assistance and space, and Daryl Domman for providing *V. cholerae* alignments used as a background phylogenetic dataset. We also thank the original authors of the sequences used in this background dataset (references provided in Appendix). Funding for this project was provided by Bill and Melinda Gates Foundation OPP1195157.

## Notes

### Competing Interest Statement

The authors have declared no competing interest.

### Author Declarations

The Johns Hopkins Bloomberg School of Public Health Institutional Review Board waived ethical approval for this study.

